# Near-Infrared Imaging of Indocyanine Green Identifies Novel Routes of Lymphatic Drainage from Metacarpophalangeal Joints in Healthy Human Hands

**DOI:** 10.1101/2020.11.04.20225995

**Authors:** H. Mark Kenney, Gregory Dieudonne, Seonghwan Yee, Jeffrey H. Maki, Ronald W. Wood, Edward M. Schwarz, Christopher T. Ritchlin, Homaira Rahimi

## Abstract

**Background:** Collecting lymphatic vessel (CLV) dysfunction has been implicated in various diseases, including rheumatoid arthritis (RA). Previous studies in the tumor necrosis factor-transgenic (TNF-Tg) mouse model of inflammatory-erosive arthritis have demonstrated reduced joint-draining CLV contractility that correlates with arthritic severity. Clinically, RA patients with active hand arthritis exhibit significantly reduced lymphatic clearance of the web spaces adjacent to the metacarpophalangeal (MCP) joints and a reduction in total and basilic-associated CLVs on the dorsal surface of the hand by dynamic near-infrared (NIR) imaging of indocyanine green (ICG). In this pilot study, we assessed direct lymphatic drainage from MCP joints, and aimed to visualize the total lymphatic anatomy using novel dual-agent relaxation contrast magnetic resonance lymphography (DARC-MRL) in the upper extremity of healthy human subjects.

**Methods:** Two healthy male subjects participated in the study. We performed NIR imaging following intra-dermal web space and intra-articular MCP joint injections of ICG to visualize the CLV anatomy on the dorsal surface of the hand and antecubital fossa. Subsequently, we performed conventional or DARC-MRL after intra-dermal web space and intra-articular MCP joint injections of gadolinium to evaluate the total lymphatic anatomy of the upper extremity and compare with NIR-ICG imaging.

**Results:** NIR-ICG imaging demonstrated that web space and MCP lymphatics drain via distinct CLV tributaries. Web space draining CLVs tended to be associated with the cephalic side of the antecubital fossa, while MCP draining CLVs were localized to the basilic side of the forearm. The DARC-MRL methods used in this study did not adequately nullify the contrast in the blood vessels, and limited gadolinium-filled CLVs could be identified.

**Conclusion:** Through the use of NIR-ICG imaging, we found that MCP joints predominantly drain into basilic CLVs in the forearm, which may explain the reduction in basilic-associated CLVs in the hands of RA patients. In healthy subjects, current DARC-MRL techniques have limited ability in identifying lymphatic structures and further refinement in this technique is necessary.

## Introduction

The lymphatic system regulates local tissue fluid volumes and serves as a conduit for immune cells.^1-3^ Within the tissue, lymphatic capillaries are the first vessels to absorb interstitial fluid, which then flows into larger collecting lymphatic vessels (CLVs). CLVs are surrounded by lymphatic muscle cells that mediate spontaneous and high-amplitude contractions to unidirectionally shuttle lymph through a series of lymph nodes ultimately draining into the cardiovascular system at the junction of the internal jugular and subclavian veins.^1, 2^ Proper lymphatic fluid flow is essential for homeostasis, and our understanding of CLV dysfunction in various disease states has evolved considerably in recent years.^4-6^

One specific example of lymphatic dysfunction in disease is rheumatoid arthritis (RA). Although RA is considered primarily an autoimmune disease, growing evidence supports the contribution of epigenetic and non-adaptive mechanisms associated with the onset of arthritis.^7^ For instance, despite the fact that rheumatoid factor (RF) and anti-citrullinated protein antibodies (ACPAs) are effective biomarkers for RA, the involvement of autoantibodies is neither necessary (seronegative RA) nor sufficient (autoantibodies can precede joint damage by many years) to cause RA.^8^ Thus, additional mechanisms are likely functioning in RA pathogenesis to promote inflammatory-erosive arthritis alongside immune dysfunction.^7^ Given that RA affects 0.5 – 1% of the population,^7, 9^ and the pathologic mechanisms that mediate seronegative RA are poorly understood,^10^ research is needed to develop more targeted treatments and effective clinical biomarkers of disease severity and treatment response.

Tumor necrosis factor transgenic (TNF-Tg) mouse models of spontaneous, non-antibody mediated inflammatory-erosive arthritis were the first to provide evidence of lymphatic dysfunction correlated with disease severity.^5, 6^ The TNF-Tg mice demonstrate a progressive reduction in joint-draining popliteal CLV contractility and decreased lymphatic clearance as measured by near-infrared (NIR) imaging of indocyanine green (ICG).^5, 11-16^ Clinically, RA subjects with active hand disease were similarly shown by NIR-ICG imaging to have significantly reduced lymphatic clearance of the web spaces adjacent to the metacarpophalangeal (MCP) joints and a decreased number of CLVs on the dorsal surface of the hand compared to healthy controls.^17^ Interestingly, the loss of CLVs in RA patients tended to be localized to the basilic-associated CLVs on the surface of the hand.^17^ These first-in-man studies validated the novel paradigm of lymphatic dysfunction as a potential component of RA pathogenesis that warranted further investigation by directly assessing lymphatic drainage of MCP joints. To study MCP joint-draining lymphatics in RA, we must first evaluate the feasibility of directly assessing MCP joint-draining CLVs in longitudinal imaging studies. Most importantly, we must also determine if CLVs identified by NIR-ICG imaging serve as a biomarker of total lymphatic tributaries in the human hand and forearm.

Herein, we performed a pilot study of two healthy subjects comparing the CLV anatomy on the dorsal surface of the hand and the antecubital fossa between intra-dermal web space versus intra-articular MCP joint injections of ICG visualized by NIR imaging. The primary disadvantage of NIR-ICG imaging is the shallow depth in tissue penetration, which limits visualization of CLVs to those within 1 – 2 centimeters below the skin surface.^18^ Thus, in order to interrogate the deep CLVs that may drain the web space and/or MCP joints, we performed magnetic resonance lymphography (MRL) in these same subjects. We performed MRL using a combination of gadolinium contrast concurrent with intravenous iron administration to nullify the contrast signal in the blood vessels and perform isolated lymphatic imaging. We evaluated dual-agent contrast enhanced MRL in imaging healthy lymphatic vessels using a conventional fast spoiled gradient-echo (FSPGR) sequence or the application of the Dixon method of uniform fat suppression, with which satisfying success has been established and termed dual-agent relaxation contrast (DARC)-MRL. ^19, 20^ The DARC-MRL method has successfully visualized the tortuous and dilated CLVs in patients with lymphedema.^20^ In this study, we are the first to attempt DARC-MRL imaging of healthy CLVs to determine if current DARC-MRL parameters are sufficient for this population and to validate CLV anatomy as imaged with NIR-ICG techniques.

## Methods

### Clinical Trial Design

All of the experiments were approved by the University of Rochester IRB and FDA (NCT04046146). A total of two healthy male subjects were involved in this pilot study. Both subjects were provided informed consent and were 18 years of age or older. Subjects were excluded based on the following criteria: 1) active systemic disorders or inflammatory conditions (i.e. chronic infections with hepatitis B, hepatitis C, or HIV) that would confound the study results; 2) known sensitivity to iodine because of residual iodide in indocyanine green; 3) known sensitivity to gadolinium; and 4) any history of kidney disease (including kidney tumor, single kidney, or kidney stones) resulting in an estimated glomerular filtration rate (eGFR) < 60mL/minute per 1.73m^2^ within 1 year of the study visit.

We conducted lymphatic imaging of the hands and forearms at four visits using different methods of imaging and routes of contrast administration: 1) NIR imaging following intra-dermal ICG web space injections (visit 1), 2) MRL following intra-dermal gadolinium-diethylenetriamine pentaacetic acid (Gd; Gadavist [gadobutrol], Bayer; Leverkusen, Germany) web space injections (visit 2; > 1 week after visit 1), 3) NIR imaging following intra-articular ICG MCP joint injections (visit 3; > 8 weeks after visit 1), and 4) MRL following intra-articular Gd MCP joint injections (visit 4; > 1 week after visit 3). The first subject completed all 4 visits, while the second subject completed the first 3 visits. Up to 10 subjects were planned to be enrolled in the study, but the experiments (including Subject 2, visit 4) were cancelled due to regulations associated with the COVID-19 pandemic at the University of Rochester where this study was conducted.

The results of this study are primarily descriptive as the study was not powered for quantitative analysis, although the trends in the lymphatic anatomy between the two subjects were consistent. All data can be made available upon reasonable request.

### Near-Infrared Indocyanine Green (NIR-ICG) Imaging

For the NIR-ICG imaging, 0.1mL of 100μM ICG was injected intra-dermally into the web spaces or intra-articularly into the MCP joints. Web space injections were performed in both hands, while the MCP joint injections were administered in the non-dominant hand to limit the potential for pain with the intra-articular injections (although both subjects reported reduced pain with the intra-articular injections compared to the web space injections). The NIR-ICG imaging was performed on a custom-built FDA approved system, and the details for the hand imaging are described previously.^17^ Briefly, after the hand imaging was complete, antecubital fossa imaging was performed while the patient was sitting with the arm supinated and the elbow in slight flexion. Since the elbow is flexed, we did not perform the imaging using a standard distance from the lip of the camera to the antecubital fossa, as was previously described for the hand (60 ± 1cm to 3^rd^ MCP joint). For the antecubital fossa, the height of the camera was not adjusted from the distance determined for the hand. The delivered intensity was assessed by a power meter (ThorLabs USB Power Meter, Cat# PM16-121) held at the center of the antecubital fossa prior to imaging as to not exceed the FDA approved excitation intensity threshold of 1.8mW/cm^2^, although a standard illumination intensity was not used as in the hand (950 ± 20μW/cm^2^). The hands and antecubital fossas were imaged for 11 minutes each with the following imaging parameters: 500 ms camera shutter speed, no binning, 400 gain, and f/4-5.6 aperture. The individual frames were imported into ImageJ, and stacked videos were generated to visualize real-time lymphatic contractions and confirm lymphatic anatomy. Given the variability in signal intensity between subjects and routes of contrast administration, images are depicted to highlight the anatomy with manually adjusted thresholds and max intensity projections of multiple frames. The original images can be provided upon reasonable request.

### Magnetic Resonance Lymphography (MRL)

For the web space visits, we captured the MRL using two variants of DARC-MRL,^19, 20^ which uses intravascular ferumoxytol to null any gadolinium that enters the vascular system: 1) fast spoiled gradient-echo with lengthened TE (FSPGR; Subject 1, Fig. 4) and 2) the published DARC-MRL using the Dixon method for uniform fat suppression (DARC-MRL; Subject 2, Supplementary Fig. 2).^19, 20^ We also used this latter method for the MCP joint injections in Subject 1 (Fig. 5). DARC-MRL has been previously shown to successfully acquire pure lymphatic images without vascular contamination,^19, 20^ and was thus considered the most appropriate method to visualize healthy lymphatics. The FSPGR sequence is a typical method of vascular imaging, and our use of this sequence is attributed to the fact that the required lengthened TE for vascular suppression could not be dialed in using the commercially available Dixon imaging sequence. We utilized similar dual-agent contrast enhancement for both approaches in this study.

To capture the MRL imaging, subjects received intravenous ferumoxytol (iron) diluted in normal saline to 60 mL total volume at 5 mg iron/kg body weight at a rate of 0.1 mL/sec. Subjects were then administered 0.5 mL of Gd diluted in saline. For the intra-dermal web space injections, we used 1:15 dilution of Gd: saline in Subject 1 and 1:20 dilution of Gd: saline in Subject 2. For the intra-dermal web space injections, the dilution factor proved to be too concentrated for visualization (Fig. 4 [Subject 1; FSPGR] and Supplementary Fig. 2 [Subject 2; DARC-MRL]). In the intra-articular MCP joint injections for Subject 1, we similarly used 1:15 dilution of Gd: saline for consistency with the web space visit in this subject, but this time demonstrated robust contrast signal in the MCP joints using the Dixon method (Fig. 5; DARC-MRL). As in the NIR-ICG imaging, Gd was administered into both hands for the web space injections, while the Gd was only injected into the MCP joints of the non-dominant hand. Subjects were placed on their side in the MRI scanner either in “prayer” (both palms together) for simultaneous imaging of both upper extremities following intra-dermal injection or “superman” (one arm extended) for imaging of the dominant upper extremity following intra-articular injection. For the web space visit for Subject 1 (Fig. 4), we collected the MRI sequences after iron and Gd had been administered. In the web space visit for Subject 2 (Supplementary Fig. 2) and the MCP joint visit for Subject 1 (Fig. 5), “scout images” were acquired following iron administration and before Gd injections, which was useful for identifying the nulling parameters (i.e. TE) of the blood vasculature.

The subjects were imaged for about 1-hour at regular intervals using the parameters promising to yield the greatest nullification.^20^ For the web space visit of Subject 1 using the FSPGR sequence with a TE of 5.68 (Fig. 4), we did not achieve nullification of MRI signal in the blood vasculature, and thus we opted to analyze these images using a subtraction technique. This method subtracts the initial time point from later time points to assess changes in MRI signal in the vasculature. Since the iron is administered intravenously and is homogenously distributed throughout the blood, the intravascular signal should not substantially change for the duration of the imaging and thus not register post-subtraction, assuming very little subject motion during imaging. However, as Gd was administered intra-dermally and continually drained by the lymphatics, any change in signal would theoretically represent Gd in the subtraction sequences. With this logic, we segmented the blood and lymphatic vasculature following Gd web space injections. In the web space visit of Subject 2 (Supplementary Fig. 2), we collected an initial iron scout image (before Gd administration) followed by DARC-MRL imaging,^19, 20^ and thus present the original sequences. Similarly for the MCP joint injections using DARC-MRL (Fig. 5) with the addition of the iron scout image and the partial vascular nulling achieved of the MRI signal at all TR and TE settings, we chose to report our findings using TE_1_/TE_2_ = 7.8/9.0 realizing somewhat shorter TEs (TE_1_/TE_2_ = 6.8/8.0) were previously shown to be more than sufficient to null the blood vessels.^20^ A standard clinical MRI scanner (3 Tesla GE Discovery RM750W) within the Department of Imaging Sciences at the University of Rochester Medical Center was used for the DARC-MRL.

## Results

### Dynamic NIR-ICG Imaging Reveals Distinct Lymphatic Vessels Draining the MCP Joint versus the Adjacent Web Space

To evaluate the anatomy of the CLVs draining the web space, we performed intra-dermal injections of ICG into the web spaces adjacent to the MCP joints and monitored ICG drainage through the CLVs on the dorsal surface of the hand (Figs. 1A [Subject 1] & 2A [Subject 2]) and the antecubital fossa (Fig. 1A’ [Subject 1] and 2A’ [Subject 2]) by NIR-ICG imaging. At least 8-weeks after the administration of intra-dermal web space ICG, the same healthy subjects returned for intra-articular injections of ICG in the MCP joints for direct comparison of the CLV anatomy draining the web spaces versus the MCP joints (Figs. 1B [Subject 1] & 2B [Subject 2], dorsal hand; Figs. 1B’ [Subject 1] and 2B’ [Subject 2], antecubital fossa). In the NIR-ICG imaging, note the bright white ICG^+^ CLVs (green arrows) adjacent to dark blood vessels (red arrows, same vessels identified for orientation) (Figs. 1 & 2). CLVs were validated by assessing bolus flow of ICG through the vessels in dynamic videos generated from the NIR-ICG imaging (Supplementary Videos 1 – 7). ICG^+^ CLVs draining the web space visualized on the dorsal surface of the hand of Subject 1 (Fig. 1A, green arrows) were completely absent following direct MCP joint injections as no CLVs were noted on the dorsal surface of the hand in Subject 1 (Fig. 1B, white arrows). Similarly, Subject 2 demonstrated a considerable number of CLVs in the hand following web space injections (Fig. 2A, green arrows), and a majority of these CLVs were absent after MCP joint injections (Fig. 2B, white arrow). However, unlike Subject 1, there were a couple ICG^+^ CLVs noted on the surface of the hand in Subject 2 that were potentially common between the web space (Fig. 2A) and MCP (Fig. 2B) lymphatic drainage (dashed green arrows).

**Figure 1.**
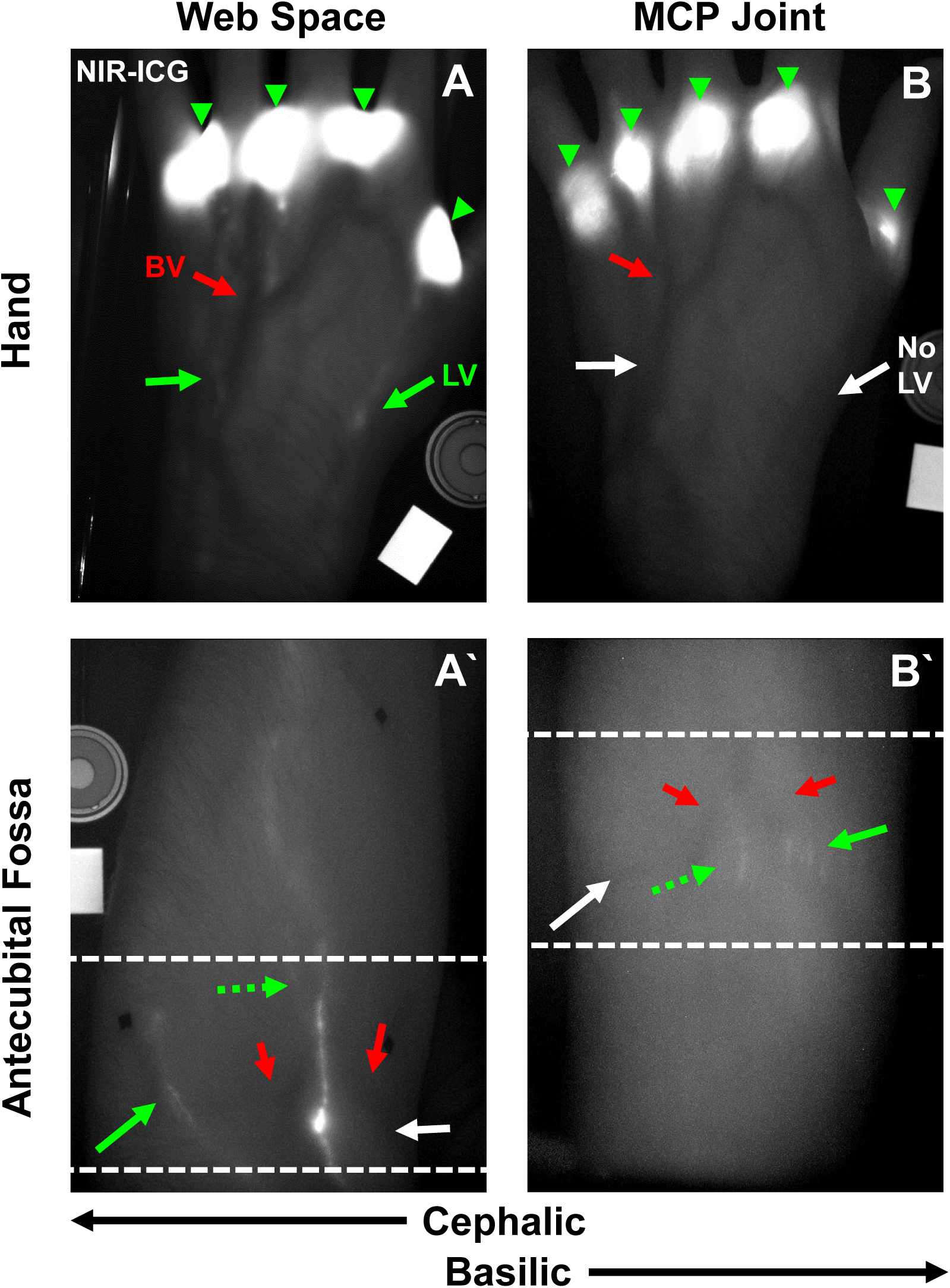
Dynamic NIR-ICG imaging reveals distinct CLVs draining the MCP joints versus the adjacent web spaces in Subject 1. NIR-ICG imaging shows ICG + CLVs (white vessels, green arrows) and adjacent superficial BVs (dark vessels, red arrows) in the hand **(A)** and antecubital fossa (**A’**, region defined by dashed white lines) following intra-dermal ICG web space injections **(A**, 4 green arrowheads). Notably in the antecubital fossa following web space injections, ICG+ CLVs are present on the cephalic side of the forearm **(A**’, green arrow). In contrast, intra-articular MCP joint injections of ICG (**B**, 5 green arrowheads) show no identifiable CLVs (white arrows) in the hand **(B)** and different draining CLVs in the antecubital fossa (**B’**, dashed lines) of the same subject. While no ICG+ CLVs were identified on the cephalic side of the forearm following MCP joint injections (white arrow), the ICG+ CLVs were localized to the basilic side of the forearm **(B**’, green arrow) that were absent following web space injections **(A**’, white arrow). A potential common CLV (dashed green arrows) from both the web space and MCP joints is noted running near the center of the antecubital fossa at the divergence of the cephalic and basilic veins (red arrows) **(A’ web space; B’ MCP joints)**.

**Figure 2.**
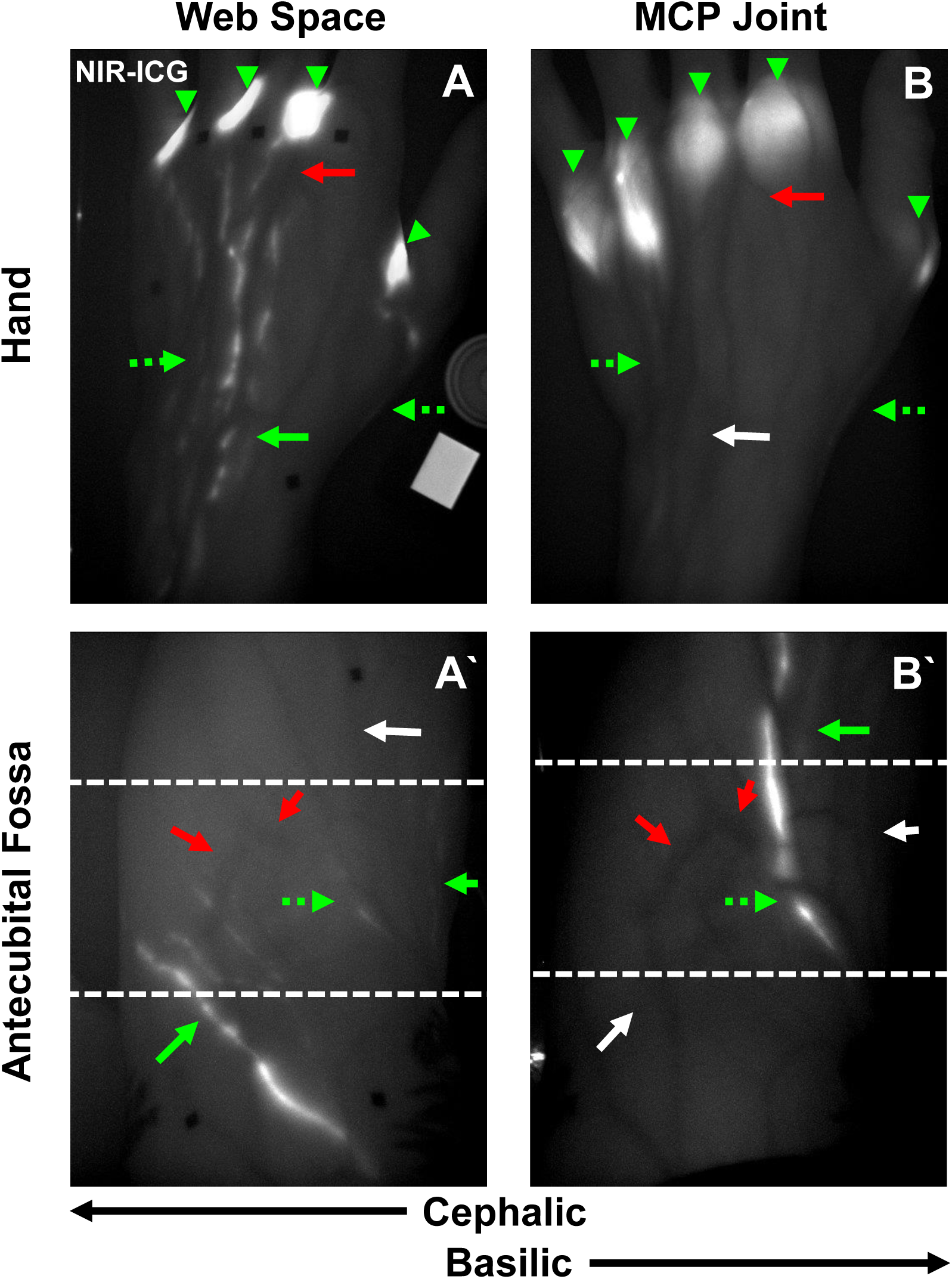
Dynamic NIR-ICG imaging confirms distinct CLVs draining the MCP joints versus the adjacent web spaces in Subject 2. NIR-ICG imaging shows ICG + CLVs (white vessels, green arrows) and adjacent superficial BVs (dark vessels, red arrows) in the hand **(A)** and antecubital fossa (**A’**, region defined by dashed white lines) following intra-dermal ICG web space injections **(A**, 4 green arrowheads). Notably in the antecubital fossa following web space injections, ICG+ CLVs are present on the cephalic side of the forearm **(A**’, green arrows). In contrast, intra-articular MCP joint injections of ICG (**B**, 5 green arrowheads) show relatively limited identifiable ICG+ CLVs (white arrow) in the hand, yet a couple ICG + CLVs are present that are potentially common to those draining the web space **(A, B**, dashed green arrows). As in Subject 1, there were distinct draining CLVs noted in the antecubital fossa (**B’**, dashed white lines) of the same subject. While no ICG+ CLVs were identified on the cephalic side of the forearm following MCP joint injections (white arrow), an ICG+ CLV was present on the basilic side of the forearm **(B**’, green arrow) that was absent following web space injections **(A**’, white arrow). However, there was also a cluster of basilic-associated CLVs present following web space injections (**A’**, green arrow) that was not identified after MCP joint injections (**B’**, white arrow). As in Subject 1, a potential common CLV (dashed green arrows) from both the web spaces and MCP joints is noted running near the center of the antecubital fossa at the divergence of the cephalic and basilic veins (red arrows) **(A’ web space; B’ MCP joints)**.

As the ICG flowed into CLVs in the antecubital fossa (dashed white lines), two prominent ICG^+^ CLVs were noted for Subject 1 (Fig. 1A’, green arrows), one running directly between the divergence of the cephalic and basilic veins (red arrows) and the other traversing the cephalic side of the forearm, following web space injections. However, after MCP joint injections, cephalic-associated ICG^+^ CLVs were notably absent (Fig. 1B’, white arrow) and instead a cluster of basilic-associated ICG^+^ CLVs were visualized (Fig. 1B’, green arrow). For Subject 2, similar cephalic-associated ICG^+^ CLVs were present following web space injections (Fig. 2A’, green arrow) that were absent after MCP joint injections (Fig. 2B’, white arrow). However, after both web space (Fig. 2A’) and MCP joint (Fig. 2B’) injections, there were distinct basilic-associated ICG^+^ CLVs (green arrows) identified following the different routes of ICG administration in Subject 2. Interestingly, for both Subjects 1 and 2, there was a potential common CLV (dashed green arrow) between both web space (Fig. 1A’ & 2A’) and MCP joint (Fig. 1B’ & 2B’) ICG administration present in the center of the antecubital fossa near the divergence of the cephalic and basilic veins (red arrows).

Thus, for both subjects, the number of CLVs on the dorsal surface of the hand was considerably reduced following MCP joint injections compared to web space administration. In addition, at the antecubital fossa, CLVs draining the web space were more prominently localized to the cephalic aspect of the forearm, while those CLVs draining the MCP joints were localized to the basilic aspect of the forearm. Interestingly, we also identified retrograde flow of ICG from the MCP to the PIP in the 2^nd^, 3^rd^, and 4^th^ digits and the DIP in the 3^rd^ digit on the palmar surface of the hand following MCP joint injections in Subject 1 (Fig. 3). The subject reported no known injuries to these fingers. This finding was not seen in imaging of Subject 2 (Supplementary Fig. 1).

**Figure 3.**
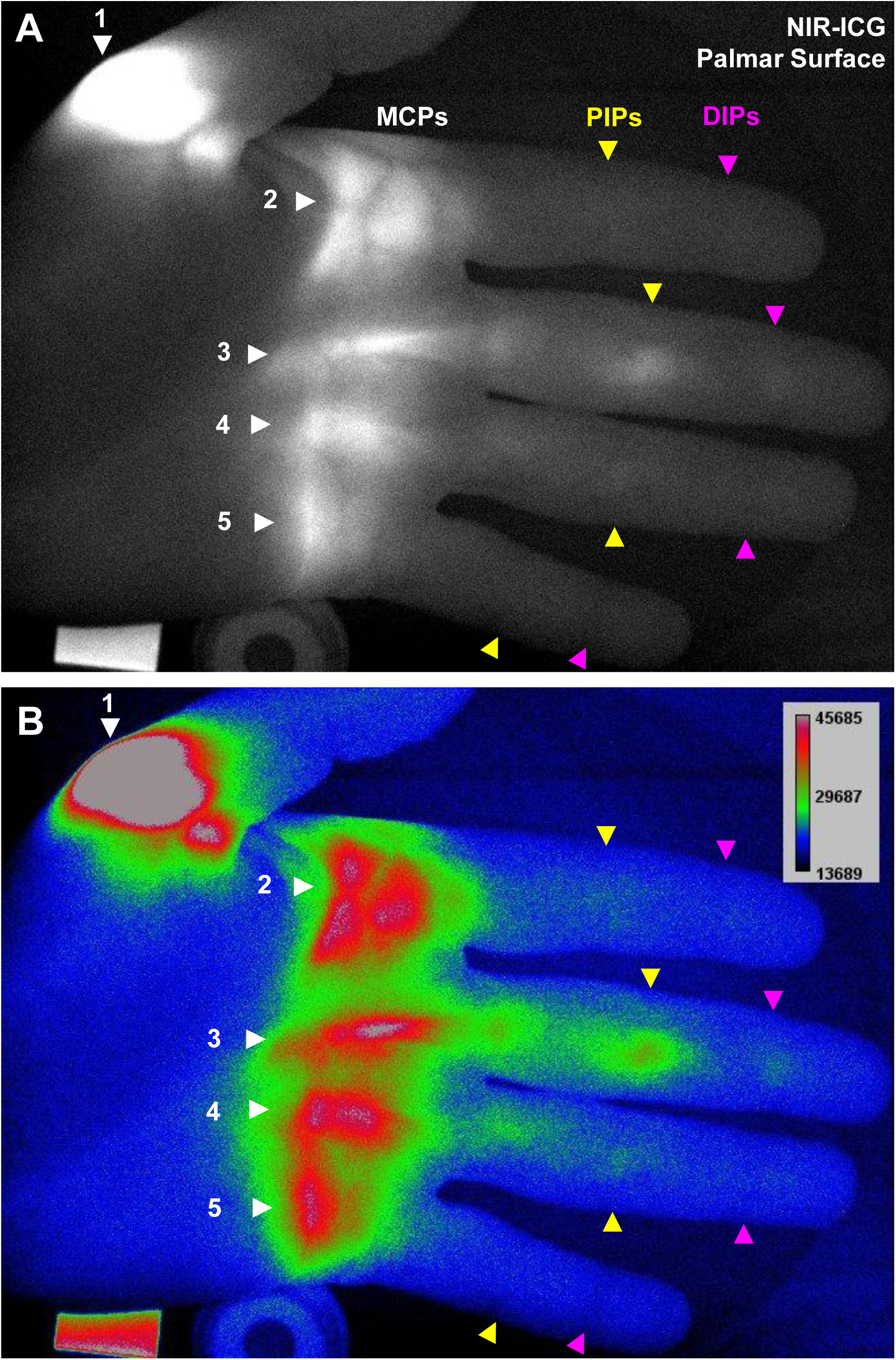
Novel retrograde ICG drainage from the MCP to the PIP and DIP joints observed via NIR-ICG imaging in Subject 1. Following intra-articular MCP joint injections of ICG, fluorescent signal was unexpectedly detected in certain distal PIP and DIP joints, most notably in the 3rd PIP (arrowheads indicate joints; white = MCPs, yellow = PIPs, pink = DIPs) **(A)** A rainbow RGB lookup table was used to visualize subtle changes in fluorescent intensity, and ICG signal was detected in the 2^nd^ PIP, 3^rd^ PIP, 3^rd^ DIP, and 4^th^ PIP (green/red signal), while the other joints did not show increased fluorescent intensity (blue) **(B)**.

### 3D MRL with Subtraction Technique Identifies Limited Web Space Draining Lymphatic Vessels Visualized in Dynamic 2D NIR-ICG Imaging

Given the difference in ICG^+^ CLVs between the web space and MCP joint injections, we suspected the MCPs to be draining via deep CLVs that were challenging to visualize by NIR-ICG imaging due to limited tissue penetrance.^18^ Thus, we investigated the feasibility of imaging healthy CLVs using conventional MRL or novel DARC-MRL imaging techniques.^19, 20^ At least 1-week after the intra-dermal web space injections of ICG, the same healthy subjects returned for intra-dermal web space injections of Gd and intravenous iron for conventional MRL imaging (Fig. 4, Subject 1) and DARC-MRL imaging (Supplementary Fig. 2, Subject 2). Single 2D images from the conventional MRL dataset were extracted, and an inverted image (note white background) representing a standard clinical T1-weighted colormap of the hand (Fig. 4A) and forearm (Fig. 4A’) are shown for anatomical orientation. Iron contrast in the blood vessels was visualized in the hand (Fig. 4B) and forearm (Fig. 4B’). Since iron contrast is distributed homogenously throughout the blood vasculature and given its relatively longer half-life, subtraction sequences from the initial time point would exclude all iron contrast and only include Gd contrast injected into the web space that is continuously being drained via the lymphatic vasculature. Using subtraction sequences from the initial time point imaged at TR 12.1 and TE 5.7, all Gd^+^ vessels were visualized in the hand (Fig. 4C) and forearm (Fig. 4C’). We noted the presence of a large BV in the forearm that was filled with both iron (Fig. 4B’) and Gd (Fig. 4C’), indicating limited nullification of the MRI signal in the blood vessels using conventional MRL imaging. Similarly, with the DARC-MRL imaging, we did not achieve nullification of the blood vasculature as demonstrated by unmodified (non-subtraction) imaging sequences of the vessels in the forearm of Subject 2 at TR 11.2 and TEs 7.8/9.0 (Supplementary Fig. 2).

**Figure 4.**
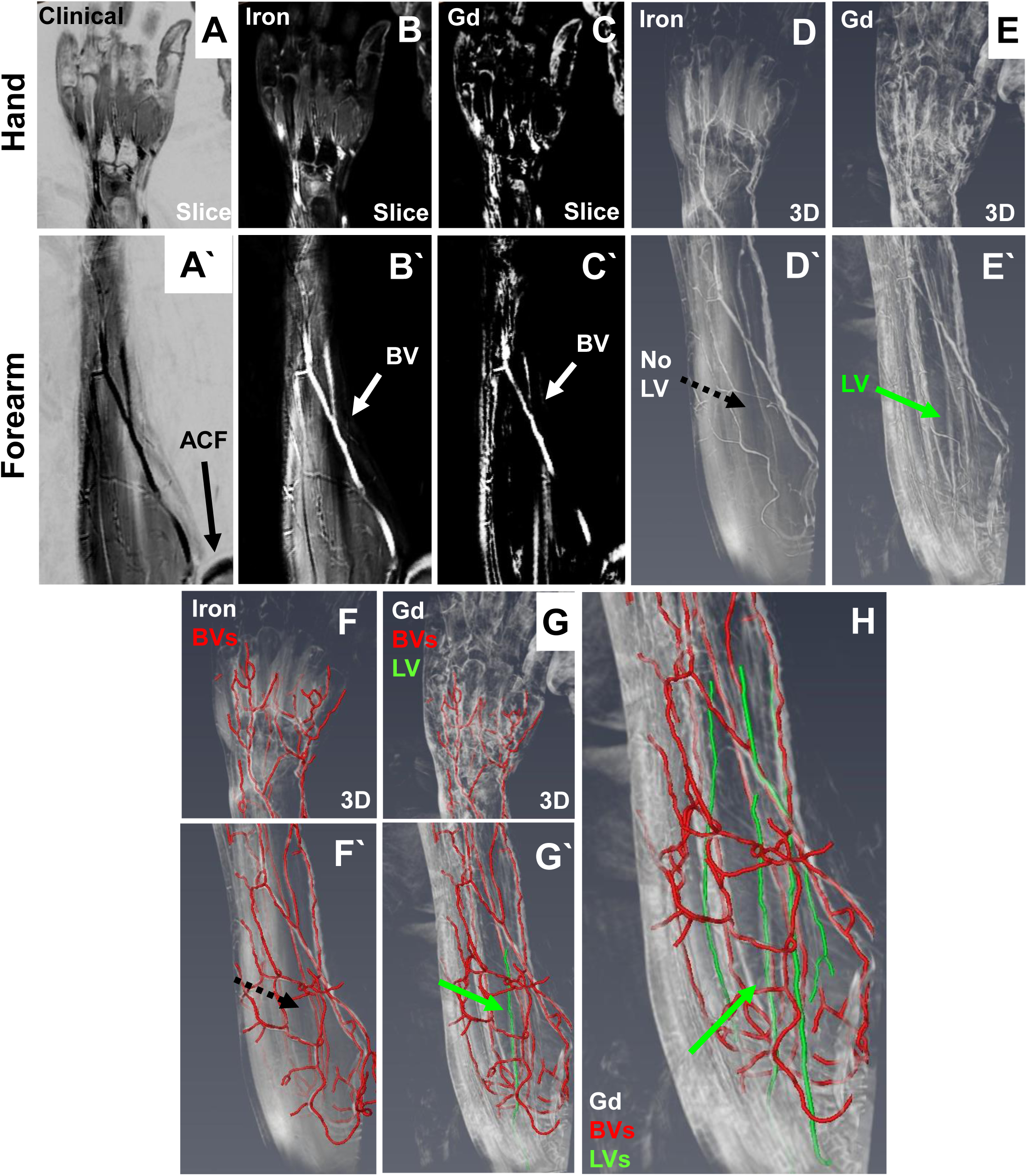
3D conventional MRL with subtraction technique identifies limited web space draining lymphatic vessels visualized in dynamic 2D NIR-ICG imaging in Subject 1. To identify the anatomical location of superficial draining LVs within the entire blood and lymphatic vasculature and assess their proportion to deep LVs that cannot be imaged by NIR, conventional MRL was performed. Intravenous iron contrast (ferumoxytol) was administered, and Gd (gadobutrol) was injected into the web spaces of the hand. Inverted representative images of the hand **(A)** and of the forearm **(A’)** in a healthy subject are shown for orientation. The initial time point shows blood vessels containing iron contrast (white) in the hand **(B)** and forearm **(B’)** before subtraction. The subtracted image partly identified the same vessels (white arrow) in the hand **(C)** and forearm **(C**’) as in the initial time point **(B, B**’). Understand that both iron and Gd are present in a large blood vessel (BV, white arrow in B’ and C’), indicating that improvements are needed to nullify iron contrast and Gd contamination in BVs using dual-contrast administration. For 3D images, volume rendering of the MRI sequences was performed using Amira software. 3D images of the iron MRI sequence demonstrating BVs (white) in the hand **(D)** and forearm **(D’)** are shown, with volume rendering of the iron subtraction sequence representing Gd filled vessels (white) in the hand **(E)** and forearm **(E’)**. Manual segmentation of the volume rendered iron filled BVs (red) in the hand **(F)** and forearm **(F’)** adjacent to segmentation of the iron subtracted and Gd-filled vessels **(red in G and G’)** allow for identification of a Gd filled vessel that is absent in the iron sequences **(dashed black arrows in D’ and F’)**, and present in the iron subtracted sequences that image Gd **(green arrows in E’ and G’)**. Based on this subtraction method, we segmented the lymphatic vasculature of the forearm **(H)**.

Using Amira computer imaging software, we performed 3D reconstructions of the conventional MRL dataset to visualize iron^+^ (Fig. 4D, D’) and Gd^+^ (Fig. 4E, E’) vessels in the upper extremity. We noted the presence of a vessel in the Gd subtraction sequence (Fig. 4E’, green arrow) that was absent in the iron sequence (Fg. 4D’, black dashed arrow). This Gd^+^/iron^-^ vessel likely represented a CLV captured in the forearm. The iron^+^ blood vessels (red) were manually segmented throughout the hand (Fig. 4F) and forearm (Fig. 4F’), and any unsegmented Gd^+^/iron^-^ vessels were segmented (green) and were considered CLVs (Fig. 4G’ demonstrates the segmentation process of a CLV [green arrow] that is absent in the iron sequences [Fig. 4F’, black dashed arrow]). No CLVs were noted in the hand (Fig. 4G), while certain CLVs were detected in the forearm (Fig. 4H, green vessels). Anatomically, one of the identified CLVs (green arrow) may have represented the common ICG^+^ CLV noted by NIR-ICG imaging (Fig. 1A’ [web spaces], 1B’ [MCP joints]; dashed green arrows). Importantly, during the web space injections of Gd, we used a concentration at which the signal was likely quenched (note lack of contrast enhancement in the web spaces, Fig. 4), and thus may explain why Gd^+^ CLVs were only seen near the antecubital fossa as the Gd was diluted by lymph volume during Gd drainage from the web spaces.

### MCP Joint-Draining Lymphatic Vessels are Not Visualized with Current DARC-MRL Methods

Similarly, we tested whether CLVs directly draining MCP joints could be visualized by DARC-MRL. At least 1-week after intra-articular injections of ICG, the same subject returned for intra-articular MCP joint injections of Gd and intravenous iron. An inverted T1-weighted image is provided for anatomical orientation of the hand (Fig. 5A) and forearm (Fig. 5A’). Iron was administered, and a scout image of iron^+^ blood vessels (red arrows) was captured at TR 9.439 and TE 5.198 of the hand (Fig. 5B) and forearm (Fig. 5B’) prior to Gd injections. After intra-articular MCP Gd injections, the injection sites were successfully visualized (Fig. 5C, black arrow heads) and partial nullification of blood vessels (red arrows) was achieved in the hand (Fig. 5C) and forearm (Fig. 5C’) with qualitatively reduced signal intensity and number of contrast-enhanced blood vessels (red arrows). A few vessels were noted directly draining the MCP joints (Fig. 5C, green arrows) that were not present in the iron scout image, and thus likely represented CLVs. Thresholding was used to segment the iron^+^ blood vessels (blue vessels, red arrows) in the hand (Fig. 5D) and forearm (Fig. 5D’) from the iron scout image (Fig. 5B, B’). Similarly, thresholding was used to segment the vessels with MRI signal (green vessels) present after the Gd MCP joint injections, where both partially quenched iron^+^ blood vessels (red arrows) and Gd^+^ CLVs (green arrows) were present in the hand (Fig. 5E; blood vessels and CLVs present) and forearm (Fig. 5E’; only blood vessels noted).

**Figure 5.**
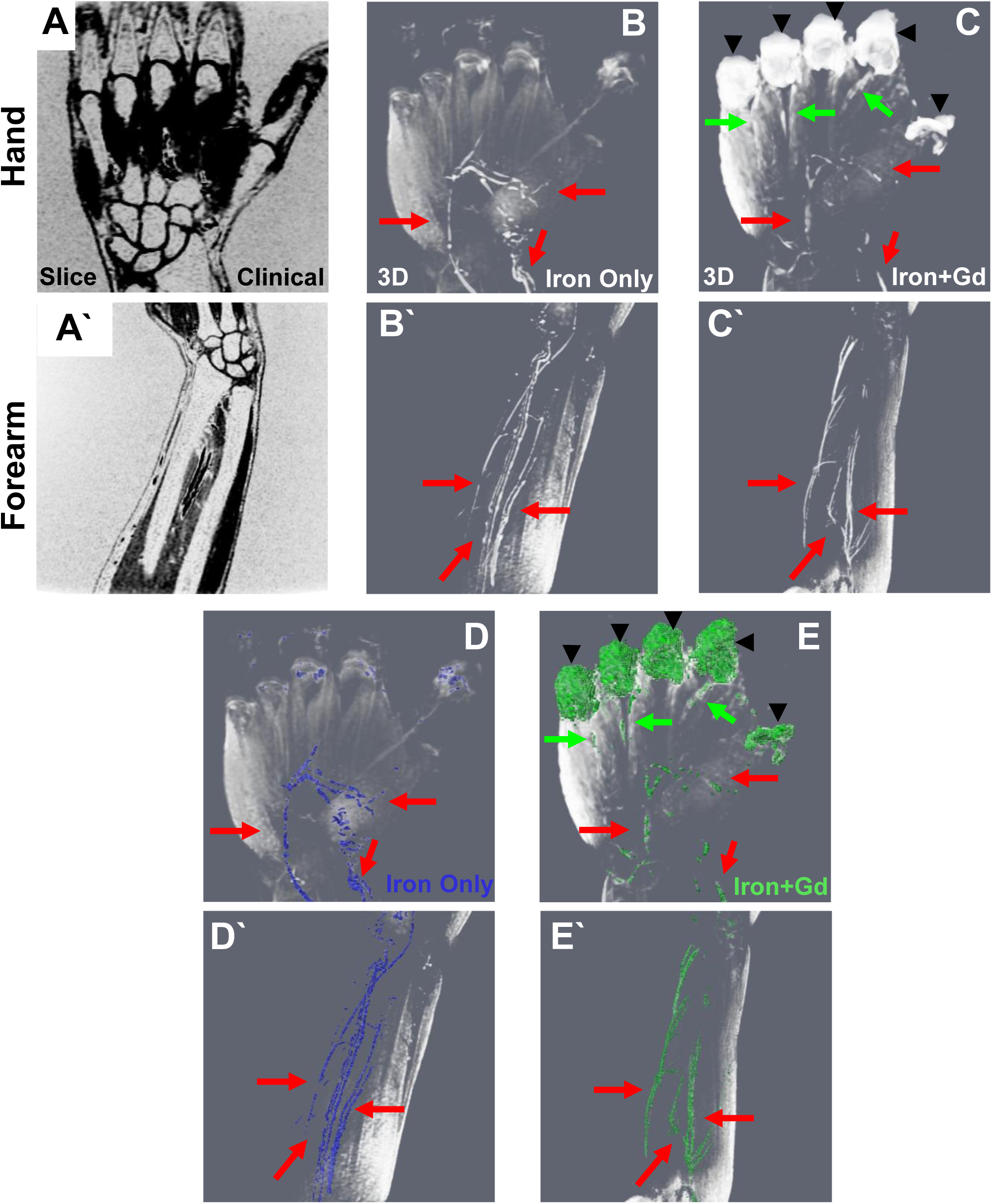
MCP joint draining lymphatic vessels are not visualized with current DARC-MRL methods in Subject 1. To assess the distinct lymphatic drainage of the MCP joints throughout the hand and forearm, DARC-MRL following gadolinium joint injections was performed on the same healthy subject. Prior to the joint injections, intravenous iron was administered and utilized as contrast enhancement to generate a scout image of the blood vasculature. Following the gadolinium joint injections, additional image sequences were captured at TE_1_/TE_2_ = 7.8/9.0; under these conditions, the iron infused blood should nullify, while the lymphatic vessels are enhanced by the gadolinium. Inverted representative images of the hand **(A)** and of the forearm **(A’)** in a healthy subject are shown for orientation. Note that the background is white, demonstrating that the sequence was inverted instead of imaged initially as a T1-weighted MRI. 3D renderings of the iron scout sequence showing the hand **(B)** and forearm **(B’)** demonstrate contrast enhanced blood vessels (red arrows). The images following the gadolinium joint injections indicate successful administration of the gadolinium into the MCP joint space (arrow heads) with contrast enhanced lymphatic vessels draining directly from the joint (green arrows) that are not visualized thereafter in the hand **(C)** or forearm **(C’)**. In addition, the blood vessels remain partially enhanced in this sequence (red arrows) demonstrating incomplete nullification using this method **(C, C’)**. Segmentation using thresholding of the contrast enhanced vasculature before **(D, D’)** and after **(E, E’)** gadolinium administration to the MCP joints shows the similar blood vessels (red arrows) and the unique lymphatic vessels (green arrows).

## Discussion

In this pilot study, we investigated the anatomy of the CLVs in the upper extremity draining the web spaces versus the MCP joints of the hands in two healthy subjects using dynamic 2D NIR-ICG imaging along with 3D conventional MRL or novel DARC-MRL techniques. To our knowledge, we are the first to demonstrate that the web space and MCP joint lymphatics of healthy human hands drain via distinct superficial CLVs by NIR-ICG imaging. Interestingly, on the dorsal surface of the hand, the number of ICG^+^ CLVs noted after intra-dermal web space ICG injections was considerably reduced following intra-articular MCP joint injections of ICG. In fact, there were no CLVs captured on the dorsal surface of the hand for Subject 1 using NIR-ICG imaging after intra-articular ICG administration into the MCPs. At the antecubital fossa, CLVs draining the web space were visualized predominantly on the cephalic side of the forearm, while CLVs from the MCP joints tended to localize to the basilic side of the forearm. This discovery generates interesting questions about the previously noted reduction in basilic-associated CLVs on the dorsal surface of the hand in RA patients with active disease^17^ because lymphatic dysfunction in RA may be related specifically to joint-draining CLVs. In support of this hypothesis, the TNF-Tg mouse model has demonstrated “expansion” and pathologic “collapse” specifically of lymph nodes efferent to joint-draining CLVs.^5,6,11,12,21,22^ Similar lymph node volumetric changes are reported in RA patients.^5,^16, 23^-25^ Interestingly, directly in the center of the forearm running between the divergence of the cephalic and basilic veins, a potentially common CLV was identified following both web space and MCP joint injections suggesting that the two tributaries may converge before the antecubital fossa. This finding provides promise that the more technically feasible assessment of web space drainage by NIR-ICG imaging has the potential to serve as a biomarker of direct MCP joint drainage by longitudinally monitoring the function of this common CLV in certain diseases, such as RA.

Given the considerable reduction in ICG^+^ CLVs on the dorsal surface of the hand, we hypothesized the MCP joints may drain predominantly via deep lymphatics not visible by NIR-ICG imaging because of the shallow depth of tissue penetrance.^18^ However, we could not rule out that the change in number of identified ICG^+^ CLVs may represent different rates of ICG absorption from the tissue, where lymphatic drainage may be more rapid from the web spaces compared to the MCP joints. While quantification of ICG clearance will be necessary to formally assess the rates of lymphatic absorption, this alternative hypothesis is unlikely to explain the lack of notable ICG^+^ CLVs on the dorsal surface of the hand given the successful transit of ICG to CLVs in the forearm at sufficient rates to be visualized by NIR-ICG imaging. Nonetheless, we attempted to image the total lymphatic vasculature in the hand and forearm using conventional or DARC-MRL. We concluded that the methods we used were insufficient for imaging the CLVs in healthy subjects. Future studies should consider safely titrating the dose of iron to achieve more efficient nullification of the Gd that is inevitably drained into the blood vasculature from the lymphatic system. While we presume the dose of iron was too low in our study to properly nullify the MRI signal in the blood vessels, we could not rule out the possibility that the dose of iron was too high and instead provided undesired contrast enhancement to the blood vasculature. In addition, we also attempted to image the entire hand and forearm in a single imaging session, which necessitated a large coil typically used for cardiac imaging. To enhance the resolution of Gd^+^ CLVs, we would consider using a higher performing coil and limiting the field of view to increase the resolution for capturing small diameter CLVs draining healthy human hands. Lastly, although the addition of a subtraction technique may theoretically assist in isolating CLVs even without complete nullification of the blood vessels in conventional MRL, we cannot rule out the possibility that structures we identified as CLVs may be a product of subtle patient movements and could be a confounding factor in this study.

Another unique finding in this study was the presence of potentially retrograde lymphatic flow in an otherwise healthy extremity. In healthy CLVs, retrograde lymphatic flow is prevented by the presence of unidirectional valves,^1-3^ and lymphatic reflux is typically considered a diagnostic feature of lymphatic pathology, such as lymphedema.^26, 27^ To our knowledge, retrograde lymphatic flow has never been noted in the extremities of healthy human subjects. However, we could not rule out the possibility that the ICG enhancement shown in the interphalangeal joints distal to the MCPs occurred by traversing other structures, such as through the flexor tendon sheaths, although the connection between the synovial space and these structures remains unclear. The lack of consistency in these findings, which only occurred in certain digits of Subject 1 and was not observed in Subject 2, warrants formal investigation of joint-derived retrograde lymphatic flow in future studies.

Overall, imaging techniques that provide detailed information about CLVs will be essential in clinically evaluating and monitoring diseases with lymphatic involvement, such as RA. As demonstrated in this work and others,^5,16,17,28,29^ NIR-ICG imaging serves as a valuable tool for longitudinally assessing CLV function in human subjects given the ability to quantify individual CLVs and their contractions despite limited tissue penetrance. Relative to MRL techniques, NIR-ICG is a much more cost- and time-effective method for clinically evaluating CLV anatomy. However, in order to validate NIR imaging following intra-dermal ICG administration as an effective biomarker of total lymphatics, we must first confirm that 1) NIR-ICG captures a majority (i.e. > 80%) of CLVs and 2) the remainder (i.e. < 20%) of deep CLVs do not demonstrate specific changes compared to those superficial CLVs identified by NIR-ICG imaging in certain diseases. While the gold-standard clinical approach of lymphoscintography is effective for assessing lymphatic drainage in certain disorders, such as lymphedema,^16^ it does not provide adequate resolution to monitor more subtle changes in CLV numbers and contractile function that have been noted by NIR-ICG imaging in both RA animal models and human subjects.^5,6,11-17,28,29^ Thus, future work continuing to utilize and validate NIR-ICG imaging as a biomarker of lymphatic dysfunction has the potential to provide major advances in clinically screening and monitoring diseases with lymphatic pathology.

## Supporting information

Video 1_Web Spaces Subject 1 Hand

Video 2_Web Spaces Subject 1 ACF

Video 3_MCP Joints Subject 1 ACF

Video 4_Web Spaces Subject 2 Hand

Video 5_Web Spaces Subject 2 ACF

Video 6_MCP Joints Subject 2 Hand

Video 7_ MCP Joints Subject 2 ACF

## Data Availability

All data can be made available upon reasonable request.

## Acknowledgements

This work was supported by research grants from the National Institutes of Health (T32 GM07356, T32 AR076950, P30 AR069655, R01 AR069000, and R01 AR056702)

## Notes

### Competing Interest Statement

The authors have declared no competing interest.

### Clinical Trial

NCT04046146

### Author Declarations

IRB University of Rochester Medical Center: STUDY00003891

### Summary of Updates

1. MRL methods updated for clarity 2. Author ORCiDs linked 3. Figures moved down on page

