## Supplementary Figures for "Near-Infrared Imaging of Indocyanine Green Identifies Novel Routes of Lymphatic Drainage from Metacarpophalangeal Joints in Healthy Human Hands"

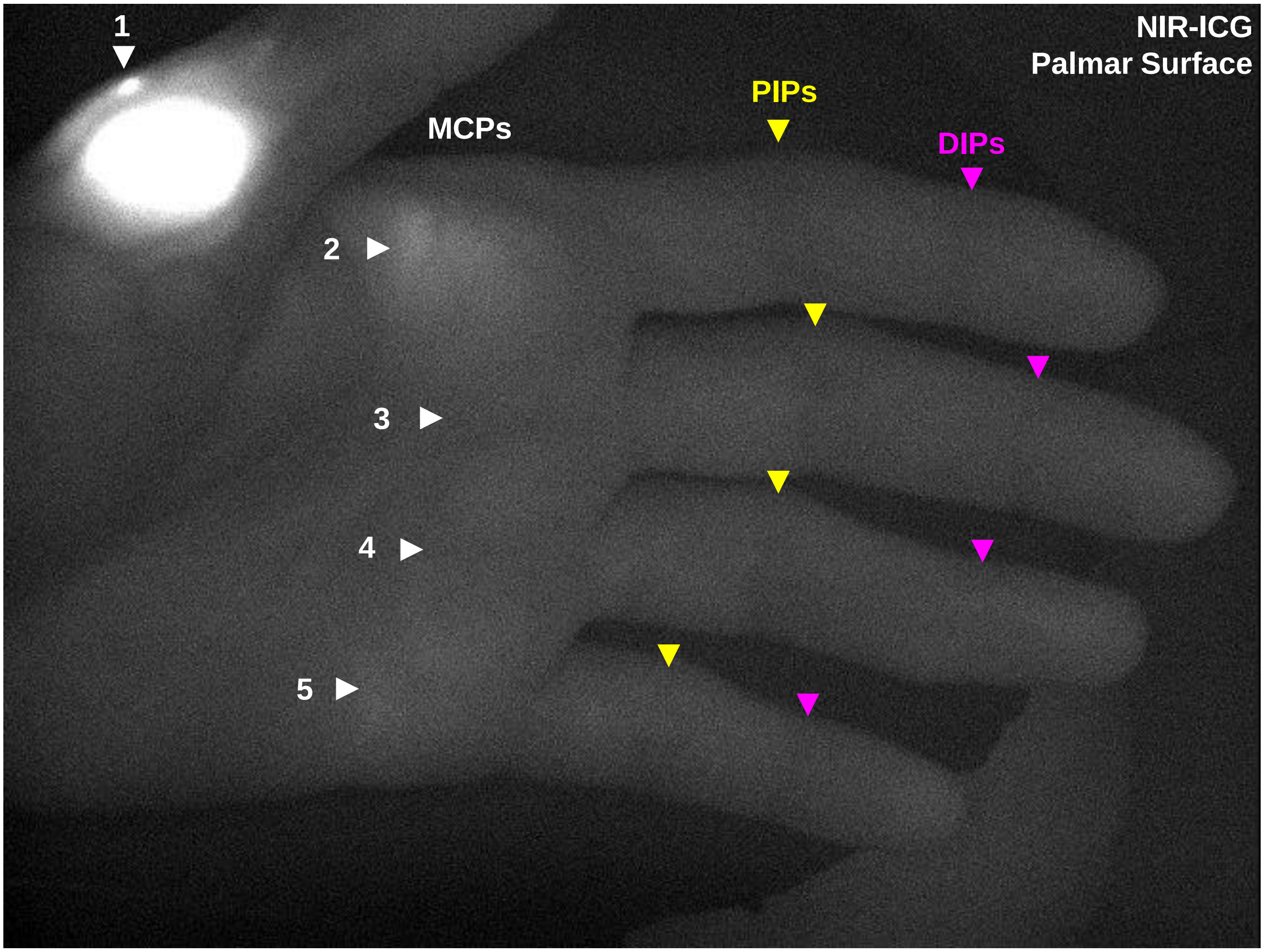


**Supplementary Figure 1. Retrograde flow of ICG was not observed in Subject 2 following intra-articular MCP joint injections by NIR imaging.** To assess the novel observation of retrograde flow of ICG from the MCP to the PIP and DIP joints in Subject 1, we similarly visualized the palmar surface of Subject 2 following MCP joint injections of ICG. Subject 2 exhibited no appreciable ICG signal in the PIPs or DIPs.

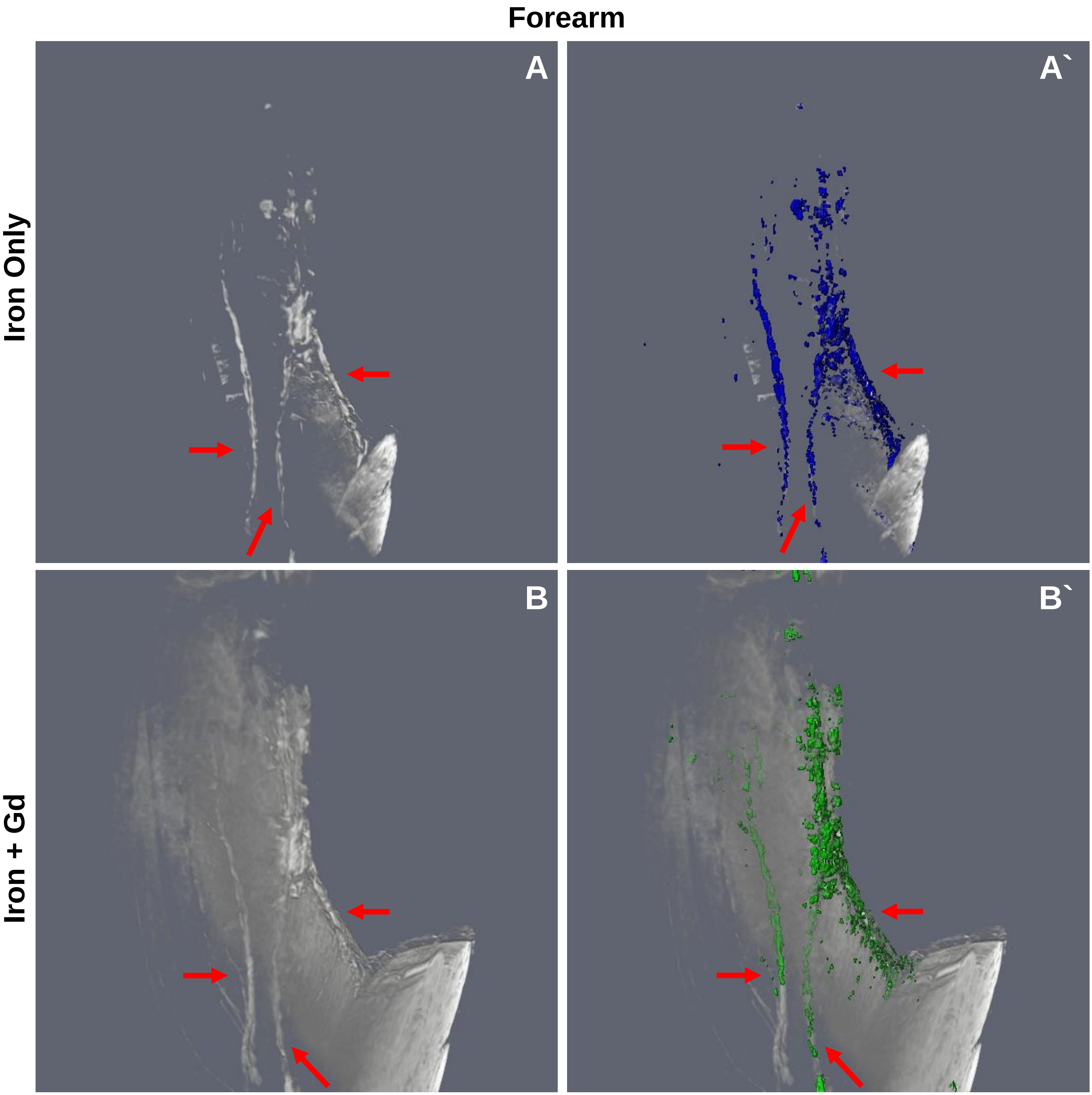


**Supplementary Figure 2. DARC-MRL did not capture lymphatic vessels draining the web spaces of healthy human hands in Subject 2.** To confirm the paucity of lymphatic vessels noted using the subtraction technique, DARC-MRL following web space injections of gadolinium was performed on the second subject. Prior to gadolinium administration, an iron infusion was delivered intravenously and used to detect the blood vasculature (red arrows) in a scout image sequenced at TR 9.439 and TE 5.198 **(A).** The blood vasculature was highlighted by segmentation using thresholding **(A`).** Following gadolinium injection into the web spaces, additional sequences were obtained at TR 12.1 and TE 8.6 demonstrating a similar vascular pattern **(B)** highlighted by segmentation using thresholding **(B`)**. The failure of achieving blood vascular nullification increased the challenge of identifying the lymphatic circulation, but a tedious review of the images did not reveal enhancement of any new vessels representing lymphatics in this healthy subject compared to the iron scout image.
